# Hospital based Multi-centric Diabetes Registry (HMDR)

**DOI:** 10.1101/2024.08.22.24311654

**Authors:** Pramila Kalra, Gurinder Mohan, Prasanta Kumar Bhattacharya, Iada Tiewsoh, K.R. Raveendra, M Ravi Kiran, Sreejith.N. Kumar, Sujoy Ghosh, S Chitra, Kaushik Pandit, Manish Chandey, Monaliza Lyngdoh, Pradip Mukhopadhyay, Richa Ghay Thaman, Smitha Jain, R Sundararaman, J Vedantha Srinivas, Veena Sreejith, Reetu Singh, Theertha Sekhar, R Mercy Elizabeth, PR Sreelakshmi

**Affiliations:** M.S. Ramaiah Medical College and Hospitals, Bengaluru, Karnataka, India; Sri Guru Ram Das University of Health Sciences, Amritsar, Punjab, India; North Eastern Indira Gandhi Regional Institute Of Health And Medical Sciences, Shillong, Meghalaya; North Eastern Indira Gandhi Regional Institute Of Health And Medical Sciences, Shillong, Meghalaya, India; Victoria Hospital (Bangalore Medical College), Bengaluru, Karnataka, India; SIMS-SRM Institutes for Medical Science, Vadapalani, Chennai, Tamil Nadu, India; Noorul Islam Institute of Medical Science and Research Foundation (NIMS Medicity), Trivandrum, Kerala, India; Institute Of Post Graduate Education and Research SSKM Hospital, Kolkata, West Bengal; Institute Of Post Graduate Education and Research SSKM Hospital, Kolkata, West Bengal, India; M.S. Ramaiah Medical College and Hospitals, Bengaluru, Karnataka

**Keywords:** Diabetes Registry, Diabetes Mellitus, Hospital Registry, Real World Data

## Abstract

Non-communicable chronic diseases, particularly Diabetes mellitus, a silent epidemic of this era, are rapidly on the rise in India. It is critical to devise strategies to ensure effective diagnosis, prevention, and treatment of the increasing disease burden. Given the lack of large scale information on Type 1 and Type 2 Diabetes mellitus, the pan-India diabetes registry is hugely pertinent to the Indian situation. The registry would be able to continuously provide data on real-world practices and standard of care for Diabetes mellitus in India. To ensure the correct blend in site selection, the centers have been chosen from different geographical regions across India with a combination of government and private sectors. All the sites will harmonize in terms of protocol, training, SOPs, and data management. This hospital-based observational cross-sectional registry will be carried out without any intervention. Patients who are known cases of Type 1 and Type 2 Diabetes mellitus visiting the outpatient departments (OPD) of all the seven Hospitals will be evaluated for eligibility and willingness to participate in future clinical trials after acquiring their informed consent or assent form (for minors). A common IT platform for entering and preserving registry data will be developed, which will enable us to establish a central database that could be utilized for future clinical trials. This will minimize the time and expense of setting up a Diabetes mellitus repository.

## Introduction

### Background/rationale

The International Diabetes Federation’s 10th edition Diabetes Atlas estimates that about 74.2 million individuals with Diabetes are currently living in India, and that figure is anticipated to reach 124.9 million by the year 2045. One in seven adults in India has Diabetes.^[1]^ Diabetes mellitus (DM) is rising in India and around the world due to longer life expectancies, altered lifestyles, and dietary habits. Among all non-communicable diseases (NCDs), the disease burden of Diabetes has increased at the highest rate (80%) between 1990 and 2016. According to published population-based surveys, the prevalence of Diabetes mellitus in India ranges from 10.2% to 36%; however, the age groups included and the methodology adopted in these surveys varied. Rapid cultural and lifestyle changes, including an aging population, increasing urbanization, dietary changes, a lack of physical activity, and unhealthy behavior, contribute to the prevalence and incidence of Diabetes mellitus. The ICMR–INDIAB study was one of the landmark articles which provided accurate and comprehensive state and national-level data on the prevalence of Diabetes in India. It was designed to cover both rural and urban areas and provide estimates for prediabetes, dyslipidemia, hypertension, obesity, and the level of glycemic control among the confirmed cases of Diabetes.^[11]^ With the aim of addressing the relative scarcity of information on youth-onset Diabetes in India, the Indian Council of Medical Research (ICMR) decided to establish the Registry of People with Diabetes with Young Age at Onset (YDR) in 2006. The major objectives of YDR are to generate information on disease patterns or types of youth-onset Diabetes, including their geographical variations within India, and to estimate the burden of diabetes complications.^[12]^ Historically, Diabetes mellitus was a disease of the affluent. However, current epidemiological research shows that Diabetes mellitus is becoming more common among middle-class and working-class people in metropolitan India.^[2, 3, 4]^ HbA1c has been suggested by the International Expert Committee and supported by the American Diabetes Association (ADA) to diagnose Diabetes in order to track the treatment of Diabetes mellitus.^[5, 6, 7]^ Numerous nations and diverse ethnic groups have adopted a cut-off value for HbA1c of 6.5% (48 mmol/mol). However, ethnicity appears to impact the cut-off values for diagnosing Diabetes mellitus.^[8, 9,10]^ There are significant data deficits in India on the distribution, trends, determinants, and outcomes of metabolic NCDs and diabetes mellitus. A registry can consistently provide information on real-world practice for monitoring and managing disease. In addition to being an essential tool for public health monitoring and research inquiry, patient registries also play a significant role in clinical research. They are a valuable resource for understanding diseases and serve society in several ways, both directly and indirectly. Many nations deploy registries to monitor the disease, develop clinician decision support systems, and understand the trends in the demographic of their populations to tailor public policies with maximum impact. This registry will facilitate gathering reliable and prospective data on type 1 and type 2 DM patients. It will assist in gathering data on disease burden, planning health care strategies, allocating resources, developing practical management guidelines, and enhancing diagnosis and treatment.

### Objectives

The primary objective of this registry is to gather information from patients with Diabetes mellitus, including demographic characteristics, their diagnoses, risk factors, complications, coexisting chronic conditions, lifestyle, and medications, to advance our knowledge of epidemiology, clinical presentation, management, and complications of Diabetes mellitus in India.

A secondary goal is to enable a baseline evaluation of the burden and treatment of individuals with Diabetes, which will provide essential data for formulating strategies in the future. This will also assist us in creating a repository of data on individuals with Diabetes mellitus that can be utilized for upcoming clinical trials and investigate the complications and comorbidities. In addition, this gives us insight into the gender and age groups most likely to develop difficulties and their relationship with one another. This will provide an opportunity to study socio–economic and cultural factors affecting access to Diabetes care and management. Finally, this will also help us facilitate the recruitment of potential participants who are interested in clinical trials.

## Methods

### Study design

This registry is an observational cross-sectional study undertaken in hospitals. Data for this registry will be gathered from various departments of hospital OPDs of 7 sites across India, which were recognized for the BIRAC project, consisting of 3 public and 4 private multi-specialty hospitals. Patients attending the hospital’s outpatient department who are known cases of Type 1 or Type 2 Diabetes Mellitus based on their medical records and aged more than a year will be enrolled in the registry after obtaining written informed consent or assent (for minors). Convenience sampling is the method used for enrolling the participant in the registry. Patients who do not have Indian citizenship, individuals with gestational diabetes mellitus, drug-induced diabetes mellitus, and secondary diabetes mellitus will not be included in the registry. Though it was ideal to choose more sites, based on the budget availability, seven sites were finalized. These seven sites include private and public hospitals as well as are from various geography across India.

Data will be collected from the eligible patients via face-to-face interviews on a printed data collection form after acquiring written informed consent or assent (for minors). Data on the demographic characteristics, medical history, family history, physical examination, personal history, complications, laboratory investigations, treatment history, and the impact of any diabetes mellitus will be captured using a Data Collection Form (DCF), which has undergone expert validation. Under demographic character, variables like gender, marital status, age, area of residence, etc., will be recorded. In medical history, the type of Diabetes Mellitus and its duration will be captured. Other variables like cardiovascular diseases, respiratory diseases, gastrointestinal diseases, and CNS diseases are also captured under medical history. The surgical history of the participant is also noted in this section. Family history of Diabetes mellitus, Hypertension, Cardio Vascular diseases, Stroke, Cancer, and Microvascular complications will also be captured. In physical examination, weight, height, BMI, waist circumference will be recorded. Personal history will include diet. It also includes the use of tobacco, cigarette, and alcohol. Variables captured under complications are Nephropathy, Ischemic Heart Diseases, Hypertension, Dyslipidaemia, Neuropathy, Retinopathy, Peripheral Arterial Disease, Diabetic foot Ulcer, Stroke, Chronic kidney disease, and infections related to or because of Diabetes Mellitus. The laboratory tests like HbA1c, serum creatinine, lipid profile, random blood sugar, fasting blood sugar and postprandial plasma glucose will be recorded. Treatment details of medication for type 1 and type 2 Diabetes Mellitus, like insulin, oral anti-diabetic medication, and Injectable diabetic medications, will be noted. If participants undergo any alternative treatment like Ayurveda, Homeopathy, etc., that will also be recorded. The impact of Diabetes Mellitus like amputation, economic burden, etc., and knowledge of patients about Diabetes Mellitus will also be captured.

Considering that the registry is not hypothetical, no power analysis and sample size calculation were performed. The data collected will be transferred into an electronic database, where it is kept securely and will remain confidential. If data is inaccurate or inconsistent, the built-in program in the data entry interface will provide an instant auto-populated query string. A unique login will be given to the Principal investigator, the data entry operator, the quality manager, the statistician, and the project manager of each site, allowing them to access the electronic web-based platform and complete the data entry, review, and approval.

To maintain uniformity across all sites with CTN diabetology, standard operating procedure(SOP) will be developed and circulated across the sites, which will include SOP on SOPs: Development, Approval and Maintaining; SOP on Roles and Responsibility of sites; SOP on Investigators and staffs; SOP on Training for investigators and staff; SOP on Pre data collection activities; SOP on Informed consent process; SOP on Interviewing techniques; SOP on data collection form; SOP on quality management; SOP on database management system; and SOP on regulatory data analysis and publication. The Registry Quality Management Plan (RQMP) serves as a management tool describing the Quality Control (QC) and Quality Assurance (QA) processes, which helps in evaluating the documentation process of all sites within the CTN Diabetology. All data obtained will be treated with confidentiality. The lead site will have access to all collaborating sites’ data to collate and analyze the entire data set, while other sites’ data is restricted through site-specific login credentials. Data discrepancies are handled by reviewing inconsistencies, investigating the cause, and resolving them with document proofs or deeming them unsolvable. Anonymized data from different patients will be aggregated and not published in a way that makes it possible to identify any specific participants. In order to prevent duplication of participants in the registry, appropriate precautions would be implemented while entering the data into the portal, such as auto-populated comments from the software.

For each participant enrolled, the software automatically generates a unique ID with the format site number followed by the serial number. In order to ensure the accuracy of the data obtained, algorithms are embedded into the software. The software is also integrated with features to check for outliers, missing data, or inconsistent data, which all can cause a disparity in the data to be generated. The software has the ability to examine consistency, incomplete fields, save features, mandatory fields, and missing fields. The mandatory fields will guarantee that all required fields are filled in. The software’s customizable dashboard, which displays the key indicators, aids in real-time tracking of registry progress.

The process of registry development has been categorized into three phases. During the preliminary phase, the protocol was designed, an electronic web-based platform and the data collecting form were developed and validated. Institutional Ethics Committee approval and registration with the Clinical Trials Registry-India (CTRI) have been done. The lead site has conducted a pilot run of the registry to determine the overall technical and operational feasibility of enrolling participants and collecting the information in the data collection form. The Ethics Committee has been notified of the necessary modifications implemented in response to the findings of the pilot run.

Before data collection, the participants will be requested to sign an informed consent and/or assent form (for minors), also indicating their willingness to be contacted in the future for clinical trials. Informed consent Form and assent form (for minors) were made accessible in English and were customized for each site. All the sites translated the informed consent and assent forms (for minors) into the local language.

All sites will maintain the hard copies of the ICF and DCF in labeled files in a secure environment to safeguard the confidentiality of the enrolled participants.

In the second phase, data collection will be done; data entry operators will be lined-up for training; the data will be transcribed into the software; the validity and accuracy of the data will be evaluated at the respective centers, and the lead site will constantly monitor the quality of the data entered. The second phase is accomplished with data collection and reporting. The third phase will involve the completion of data acquisition and the generation of the final report.

Data will be checked for duplicates and authenticated before the final analysis. While validating data, precision and accuracy will be checked in compliance with the protocol, and discrepancies such as inconsistent data, missing data, and outliers will be scrutinized. The 18.0 version of SPSS will be used to analyze the data. In accordance with the pattern of missing data, a complete case analysis will be performed without any imputation. Categorical data will be given as count and percentage, whereas continuous data will be reported as mean SD. A p-value of 0.05 or less will be deemed to be significant for all 2-tailed statistical analyses. According to the specifications, proportions, means, and standard deviation will be computed, along with a 95% confidence interval. The Chi-square test or t-test will be employed to test for differences in the various parameters, depending on the parameter type. In order to address the objectives, descriptive statistics, including univariate comparisons, will be used.

All the sites will target to contribute a maximum number of patient records with Diabetes Mellitus to the registry. Each site designated a team of investigators (Principal Investigator and Co-Investigator), Project Manager, Medical Officer, Quality Manager, Research Coordinators, Research nurse, Data entry Operator, and Biostatistician. The Investigators will be in charge of ensuring that the data input is accurate and complete.

This registry will offer the lead and participating sites vast opportunities for future clinical research. The data generated can be used to understand disease burden better, establish healthcare strategies, allocate resources, provide practical management guidelines, improve diagnosis and treatment, diminish error rates, and predict complications, prevalence, and incidence using an AI model. This registry will aid in raising diabetes awareness, understanding the existing gaps, and future policymaking.

**Figure 1:**
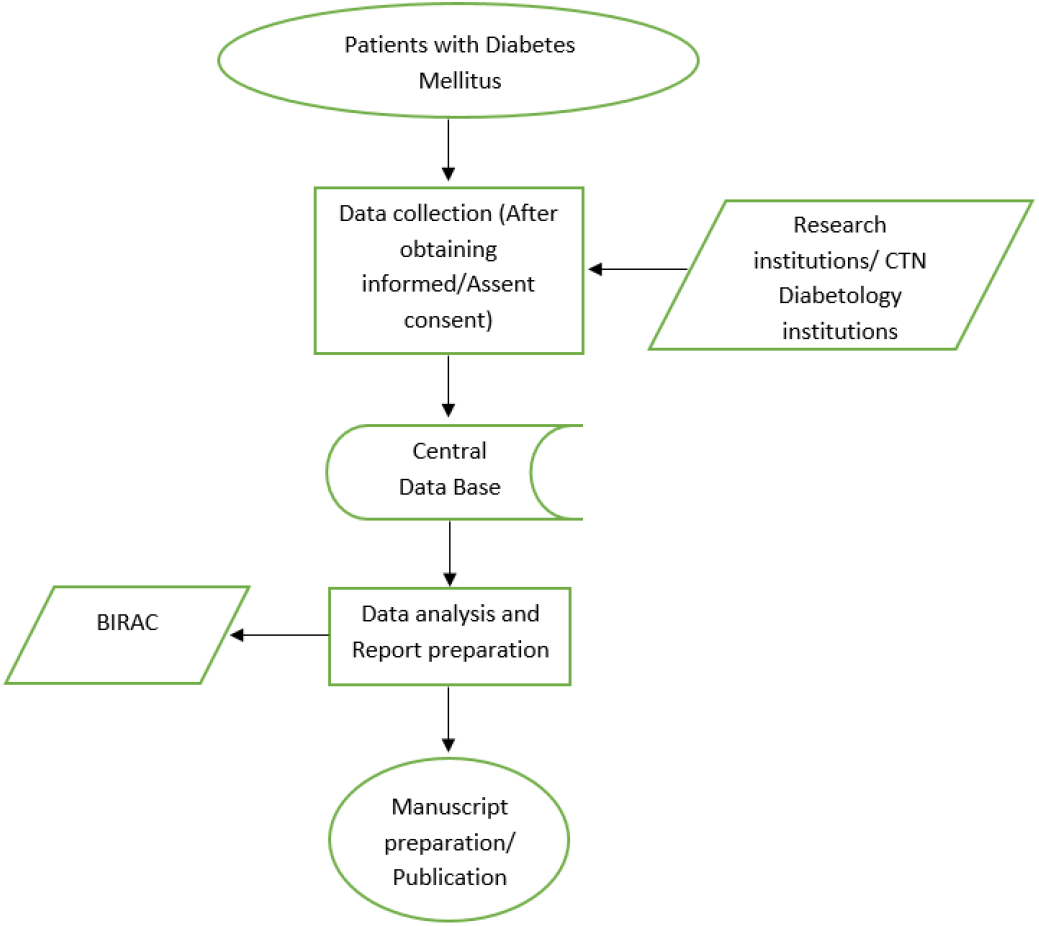
Flow chart of the study procedure

### Generalisability

With the help of this registry, we will understand the standard of care and the management techniques for Diabetes used within our country. Additionally, it provides information on how Diabetes is treated in many settings, including the public and private sectors, and for patients from various socioeconomic backgrounds. This information may be utilized to advocate for greater allocation of resources for Diabetes treatment. Further, the results could be leveraged to guide treatment recommendations and healthcare policy for those with Type 1 and Type 2 Diabetes. The registry will probably continue in the future to gather real-world practices in Diabetes care.

The database created by this registry data can be used for clinical trials in the future, reducing the cost and duration of recruitment and gathering baseline information. The tool used for the data management of the registry aids in eliminating the errors that arise during the data entry; the final dataset can be created quickly following the completion of data entry as the software has the inbuilt capacity to validate the data, which helps in real-time monitoring of the progress of registry data. First and foremost, it ensures data accuracy and prevents data duplication. In conclusion, the registry’s data may be employed to improve healthcare strategies, allocate resources, provide appropriate management directions, and enhance diagnosis and treatment. It may even be possible to forecast complications and co-morbidities using artificial intelligence or a predictive model. The registry can have an impact on legislation and continue to spread awareness about Diabetes mellitus.

## Data Availability

All data produced in the present study are available upon reasonable request to the authors

